# Prevalence of co-infection between high-risk human papillomavirus and common sexually transmitted infections in cervical specimens

**DOI:** 10.1101/2024.01.28.24301891

**Authors:** Joshua Kostera, Almedina Tursunovic, Paige Botts, Regina Galloway, April Davis, Tong Yang

**Affiliations:** Molecular Diagnostics of Abbott, Des Plaines, Illinois; Ochsner Health, New Orleans, Louisiana, USA

**Keywords:** nucleic acid amplification test, coinfection, diagnostic test, clinical laboratory, cervical cytology, human papillomavirus (HPV), sexually transmitted infection (STI)

## Abstract

**Objectives:** Evidence suggests that co-infection with sexually transmitted infection (STI) pathogens may support HPV infection persistence and cervical disease progression and neoplasia. We examined the prevalence of HPV and co-infection with other common STI pathogens in liquid-based cytology (LBC) cervical specimens, and their association with cervical disease by cytology.

**Methods:** In this retrospective, cross-sectional study, 149 randomly selected remnant cervical specimens, collected in LBC as part of routine cervical cancer screening in a large urban academic healthcare system, were tested on the Alinity m HR HPV assay and Alinity m STI assay. All specimens were processed for cytology and graded as negative for intraepithelial lesions or malignancy (NILM), atypical squamous cells of undetermined significance (ASC-US) or having any atypical cytology beyond ASC-US (≥LSIL).

**Results:** 62.4% (n=93/149) of specimens had abnormal (≥ASC-US) cytology and the remaining 37.6% (n=56/149) had NILM cytology. HR HPV was detected in 62.4% (93/149) of specimens and a single STI pathogen was detected in 11.4% (17/149) of specimens. Most specimens with a positive HPV result (73.1%; n=68/93) or positive STI result (94.1%; n=16/17) had ≥ASC-US cytology. Compared to HPV infection alone, co-infection with HPV and STI was associated with an increased prevalence of ASC-US (39.8% vs. 50.0%) and ≥LSIL cytology (33.3% vs. 41.7%). Having ≥LSIL cytology (OR= 4.1667; 95%CI: 1.6110;10.7763) or a positive STI result (OR= 1.5111; 95%CI: 0.5027;4.5420) were predictive of a positive Alinity m HR HPV assay result.

**Conclusions:** Our study confirmed an association between co-infection with HPV and STI pathogens and abnormal cytology in cervical specimens. HPV and STI co-testing may provide granular analyses of the risk of cervical disease associated with co-infections by specific HPV genotypes and STI pathogens.

**KEY MESSAGES:** *What is already known on this topic:*

- Recent studies have suggested a possible link between HPV and STI co-infection, HPV persistence in the reproductive tract, and cervical neoplasia.

*What this study adds:*

- Compared to cervical specimens positive for HPV infection alone, specimens with HPV/STI co-infection had a higher prevalence of abnormal cytology.
- Having abnormal cytology or a positive Alinity m STI assay result were predictive of a positive Alinity m HR HPV assay result.

*How this study might affect research, practice, or policy:*

- Molecular testing for high-risk HPV and common STI pathogens may reveal additional insight for predictors of cervical disease.

## INTRODUCTION

Persistent infection with high-risk human papillomavirus (HPV) is recognized as a primary cause of cervical cancer; however, there is limited information about the prevalence of co-infection with HPV and other common sexually transmitted pathogens. The cumulative lifetime risk of HPV infection is greater than 80% and is one of the most prevalent sexually transmitted infections (STIs) in the world (1). HPV infections are predominantly self-limiting and resolve within 2 years, but approximately 10% of high-risk HPV infections persist and progress to high-grade cervical disease (2). Emerging evidence suggests that concomitant infection with common STIs such as Neisseria gonorrhea (NG), Chlamydia trachomatis (CT), Trichomonas vaginalis (TV), and Mycoplasma genitalium (MG), which target the same mucosal areas of the reproductive tract as HPV, contributes to the development of cervical dysplasia and carcinogenesis in HPV-positive individuals (2,3). These STIs have also been linked to serious reproductive health complications, such as ectopic pregnancy, infertility, and pelvic inflammatory disease (3).

The mechanistic interactions between HPV and other STI pathogens have not been definitively elucidated, but several studies suggest that CT and TV may potentially favor persistent HPV infection via an inflammatory response that facilitates HPV entry into the cervical mucosa basal membrane (4, 5). Several other synergistic mechanisms influenced by STIs may also favor persistent HPV infection such as alterations in the vaginal microbiome, hormonal changes, and disruption of the cervical epithelium that increases viral load and shedding (6, 7). Algorithms for the syndromic management of STIs that use vaginal signs to predict cervical infection have unfortunately proven to be ineffective, highlighting the clinical utility of molecular testing of liquid-based cytology (LBC) specimens to aid in the diagnosis of STIs and inform patient management (8). The impact of specific or multiple HPV genotype co-infections with STIs in persistent HPV infection and pathogenesis of cervical cancer remains under investigation and is still considered controversial.

Here we present the prevalence of high-risk HPV with CT, NG, TV, and MG co-infection detected using the Alinity m HR HPV assay and Alinity m STI assay (Abbott Molecular, Des Plaines, IL) in ThinPrep® LBC cervical specimens from patients undergoing routine cervical cancer screening in an urban healthcare system. We performed an extended analysis of the age-related prevalence of HPV infection by Alinity m HR HPV assay results and cytology in our study cohort, as well as co-infection with CT, NG, TV, and MG detected by the Alinity m STI assay.

## METHODS

### Setting and Participants

Remnant non-sequential de-identified patient specimens, collected as part of the Ochsner Health routine cervical cancer screening program, were selected by the Ochsner Health molecular pathology laboratory for this study. Specimens in this study cohort were chosen to include a wide range of cytological classifications with HPV positive and negative results. Ochsner Health, based in New Orleans, is a large academic healthcare system in Louisiana. Specimens were collected in ThinPrep® LBC medium and processed for cytology. Specimens were aliquoted for molecular testing on the Alinity m HR HPV assay and the Alinity m STI assay. Patient identifiers were removed from the specimens and the study was conducted in accordance with an approved Ochsner Health Institutional Review Board (IRB) protocol number 2023.028.

### Cytology

Cytology was performed using a ThinPrep 2000 processor and specimens were graded based on the 2014 Bethesda System. Cytology results were classified as negative for intraepithelial lesions or malignancy (NILM), atypical squamous cells of undetermined significance (ASC-US), or as having any atypical cytology beyond ASC-US (≥LSIL), including low-grade squamous intraepithelial lesion (LSIL), atypical glandular cells (AGC), high-grade squamous intraepithelial lesion (HSIL), Atypical squamous cells, cannot exclude HSIL (ASC-H), and squamous cell carcinoma or adenocarcinoma. All results were reviewed by a cytotechnologist and all abnormal cases and selected negative cases (including random 10% and negative cases with previous abnormal results) were reviewed by a pathologist.

### Assays

The Alinity m HR HPV assay is a qualitative real-time PCR-based assay that simultaneously amplifies and detects genotypes HPV16, HPV18, and HPV45 and reports the 11 other HR HPV genotypes in two aggregates: Other HR HPV A (31/33/52/58) and Other HR HPV B (35/39/51/56/59/66/68). The Alinity m HR HPV assay is approved for use with ThinPrep® PreservCyt® Solution and SurePath®. The assay also includes a cellular control (CC) to ensure specimen collection adequacy along with sample extraction and amplification efficiency.

The Alinity m STI assay is a qualitative multiplex assay that simultaneously detects and differentiates CT, NG, TV, and MG nucleic acid in asymptomatic and symptomatic patient specimens. The Alinity m STI assay is cleared for endocervical swabs, vaginal (self-collected and physician-collected) and male urine for all four pathogens. ThinPrep® PreservCyt® Solution and female urine specimens are cleared for CT, NG, and TV. Rectal and oropharyngeal swabs are cleared for CT and NG. The assay also includes a cellular control to ensure specimen collection adequacy and an exogenous internal control (IC) is used to confirm the absence of PCR inhibitors in the test specimen.

### Statistical analysis

Cytology results were categorized as NILM, ASC-US, and ≥LSIL and analyzed across Alinity m HR HPV and Alinity m STI results. The analysis was further stratified by age ranges, grouped as 21-29 years, 30-44 years, and ≥45 years. Odds ratios were calculated to determine the association between an Alinity m HR HPV “Detected” result and age ≥30 years, positive Alinity m STI result (CT, NG, TV, MG), and ≥LSIL cytology. Data analyses were performed using SAS software version 9.3 or higher (SAS, Cary, NC).

## RESULTS

### Specimens

A total of 149 de-identified residual cervical clinical specimens collected in ThinPrep® were included in the study. Specimens were collected from individuals with an average age of 40.3±11.8 years. Almost half of the specimens (46%; 69/149) were collected from individuals between 30 and 44 years of age, representing the majority age group in this study; the cohort ranged from 21 to 72 years of age.

### Cytology results

Overall, 62.4% (n=93/149) of specimens had abnormal cytology (≥ASC-US) and the remaining 37.6% (n=56/149) had NILM cytology. An abnormal cytology result was more frequently found in the 30-44 age group (online supplemental table 1).

### Alinity m HR HPV results

HR HPV was detected in 62.4% (93/149) of specimens (online supplemental table 2). HPV45 was detected in 5.37% of specimens, more frequently than HPV16 (4.0%) or HPV18 (2.7%). Most HPV positive specimens were either Other HR HPV A (31/33/52/58; 18.1%) or Other HR HPV B (35/39/51/56/59/66/68; 32.2%).

The distribution of cytology by Alinity m HR HPV result and age is shown in Table 1. Of the 94 specimens with HPV positive results, 5 specimens (5.4%) had multiple HR HPV genotypes detected: 3 with HR Other A and HR Other B (1 NILM, 1 ASC-US, 1 ≥LSIL); 1 with HPV 18 and HR Other B (≥LSIL), and 1 with HPV 45 and Other HR A (≥LSIL). HPV positive results were evenly distributed by age (Table 2). The average age of individuals with a single HR HPV detected was 38.9±11.7 years and with multiple HPV detected was 35.4±11.0 years.

**Table 1.**
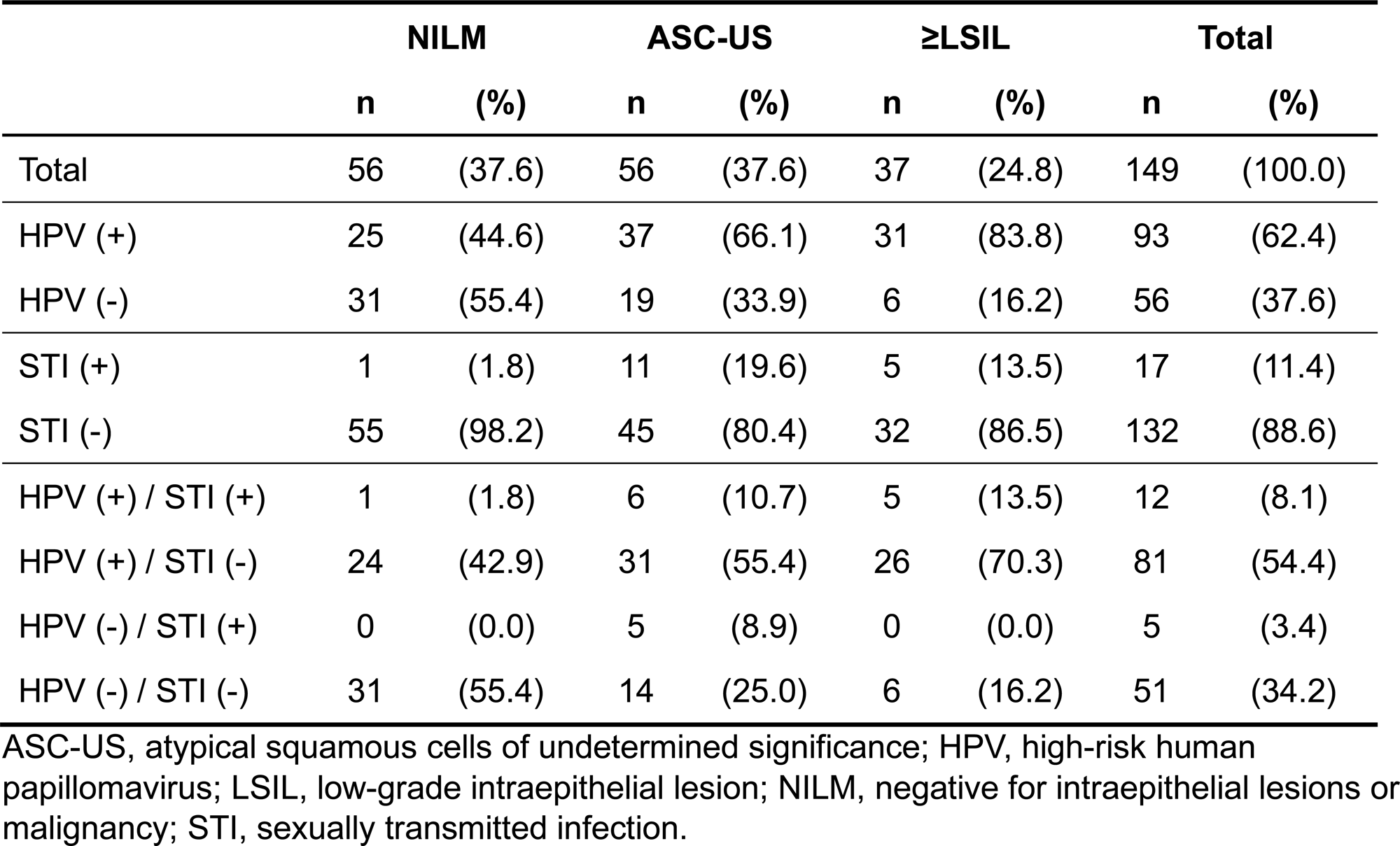
Distribution of Cytological Findings, by Alinity m HR HPV and STI Assay Results and Age Group (N=149)

| | NILM | | ASC-US | | ≥LSIL | | Total | |
| --- | --- | --- | --- | --- | --- | --- | --- | --- |
|  | n | (%) | n | (%) | n | (%) | n | (%) |
| Total | 56 | (37.6) | 56 | (37.6) | 37 | (24.8) | 149 | (100.0) |
| HPV (+) | 25 | (44.6) | 37 | (66.1) | 31 | (83.8) | 93 | (62.4) |
| HPV (-) | 31 | (55.4) | 19 | (33.9) | 6 | (16.2) | 56 | (37.6) |
| STI (+) | 1 | (1.8) | 11 | (19.6) | 5 | (13.5) | 17 | (11.4) |
| STI (-) | 55 | (98.2) | 45 | (80.4) | 32 | (86.5) | 132 | (88.6) |
| HPV (+) / STI (+) | 1 | (1.8) | 6 | (10.7) | 5 | (13.5) | 12 | (8.1) |
| HPV (+) / STI (-) | 24 | (42.9) | 31 | (55.4) | 26 | (70.3) | 81 | (54.4) |
| HPV (-) / STI (+) | 0 | (0.0) | 5 | (8.9) | 0 | (0.0) | 5 | (3.4) |
| HPV (-) / STI (-) | 31 | (55.4) | 14 | (25.0) | 6 | (16.2) | 51 | (34.2) |

ASC-US, atypical squamous cells of undetermined significance; HPV, high-risk human papillomavirus; LSIL, low-grade intraepithelial lesion; NILM, negative for intraepithelial lesions or malignancy; STI, sexually transmitted infection.

**Table 2.** Alinity m HR HPV and Alinity m STI Assay Result Distribution by Age (N=149)

|  | 21 – 29 years |  | 30 – 44 years |  | ≥45 years |  | Total |  |
| --- | --- | --- | --- | --- | --- | --- | --- | --- |
|  | n | (%) | n | (%) | n | (%) | n | (%) |
| Total | 27 | (18.1) | 69 | (46.3) | 53 | (35.6) | 149 | (100.0) |
| HPV (+) | 21 | (14.0) | 41 | (27.5) | 31 | (20.8) | 93 | (62.4) |
| HPV (-) | 6 | (4.0) | 28 | (18.8) | 22 | (14.8) | 56 | (37.6) |
| STI (+) | 7 | (4.7) | 7 | (4.7) | 3 | (2.0) | 17 | (11.4) |
| STI (-) | 20 | (13.4) | 62 | (41.6) | 50 | (33.6) | 132 | (78.5) |
| HPV (+) / STI (+) | 5 | (3.4) | 5 | (3.4) | 2 | (1.3) | 12 | (8.1) |
| HPV (+) / STI (-) | 16 | (10.7) | 36 | (24.1) | 29 | (19.5) | 81 | (54.4) |
| HPV (-) / STI (+) | 2 | (1.3) | 2 | (1.3) | 1 | (0.7) | 5 | (3.4) |
| HPV (-) / STI (-) | 4 | (2.0) | 26 | (17.4) | 21 | (14.0) | 51 | (33.6) |

HPV, high-risk human papillomavirus; STI, sexually transmitted infection.

### Alinity m STI results

Of the 149 specimens, 17 (11.4%) were positive for a single STI pathogen. No STI co-infections were detected in this cohort. The vast majority of specimens with a positive STI result (94.1%; n=16/17) had ≥ASC-US cytology (Table 1). Specimens with a positive STI result were most often from individuals ≤44 years of age (9.4%; n=14/149; Table 2). The full distribution of the 17 positive Alinity m STI assay results by STI pathogen, Alinity m HR HPV result, and cytology is shown in online supplemental table 3.

### Association of Alinity m HR HPV and STI assay results with cytology

An Alinity m HR HPV positive result was statistically significantly more closely associated with a ≥LSIL (*p* = 0.0002) cytological result than an ASC-US (*p* = 0.0226) cytological result when compared to a NILM result (Table 1). In this study 66.1% (n=37/56) of specimens with ASC-US cytology were positive for HPV; similarly, 83.8% (n=31/37) of those with ≥LSIL cytology had a positive HPV result. 44.6% (n=25/56) of those with NILM cytology were positive for HR HPV with Alinity m. Furthermore, an Alinity m STI positive result was statistically significantly more closely associated with an ASC-US (*p* = 0.0023) cytological result than a ≥LSIL (*p* = 0.0242) cytological result when compared to a NILM result.

3.4% (n=5/149) of specimens were STI positive but HPV negative, and 8.1% (n=12/149) were positive for both STI and HPV (Table 2). An Alinity m HR HPV positive result with an Alinity m STI positive result was more frequently observed and statistically significantly associated in specimens with ≥LSIL (*p* = 0.0003) cytology more so than those with ASC-US (*p* = 0.0012) cytological results when compared to NILM. Figure 1 illustrates the relationship between cytology and HPV and STI assay results. In this study cohort, an overall increasingly positive trend toward abnormal cytology was observed in specimens with an HPV positive result only and HPV/STI co-infection.

**Figure 1.**
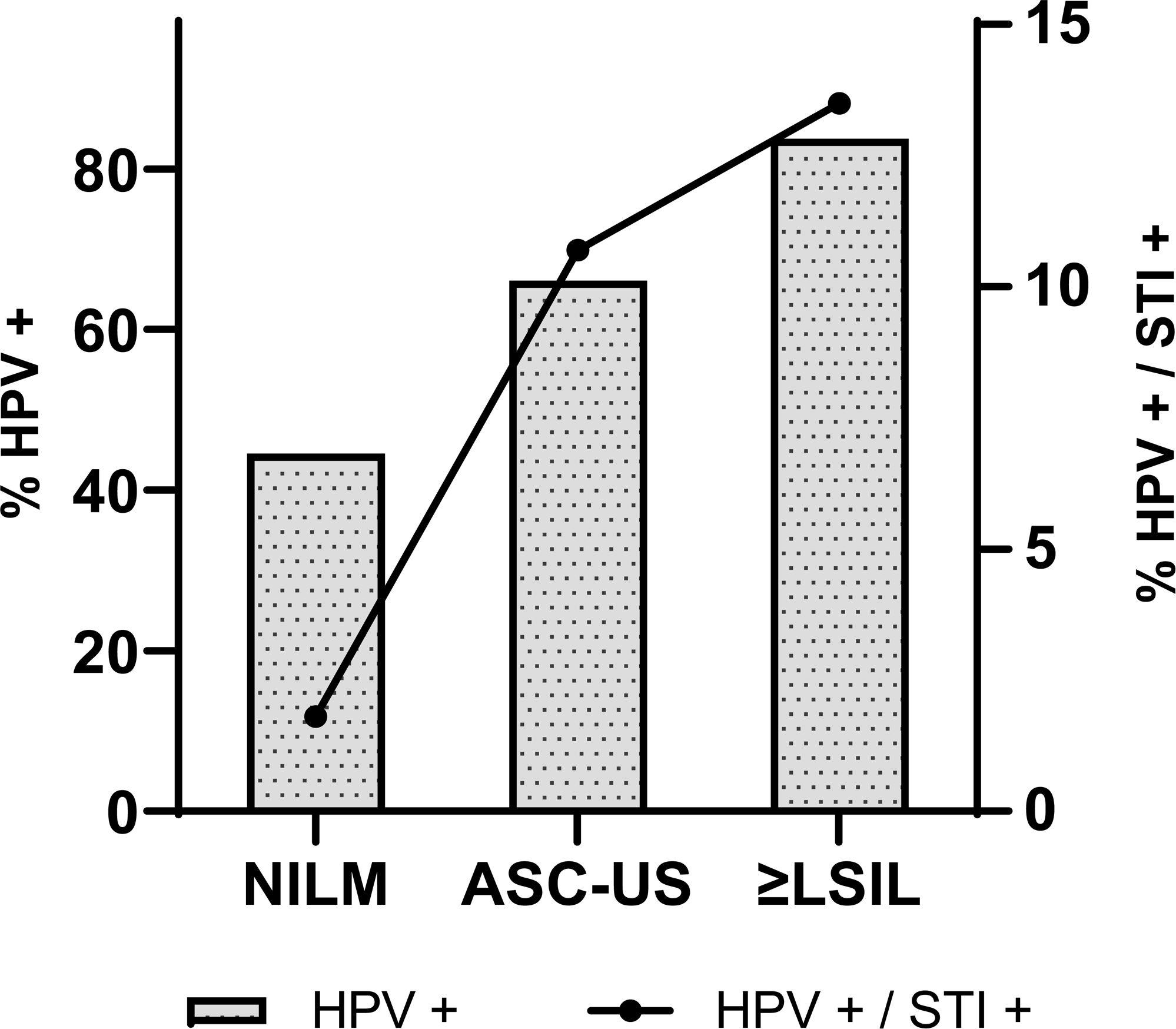
Percentage of specimens with Alinity m HR HPV and Alinity m STI positive results across cervical cytology categories.

### Predictors of a positive Alinity m HR HPV assay result

Cytology ≥LSIL (OR= 4.1667; 95%CI: 1.6110;10.7763) was most predictive of a positive Alinity m HR HPV assay result, followed by a positive STI result (OR= 1.5111; 95%CI: 0.5027;4.5420) (Figure 2). Age ≥30 years (OR= 0.4114; 95%CI: 0.1550;1.0923) was less predictive of a positive HR HPV result then either ≥LSIL cytology or a positive STI assay result.

**Figure 2.**
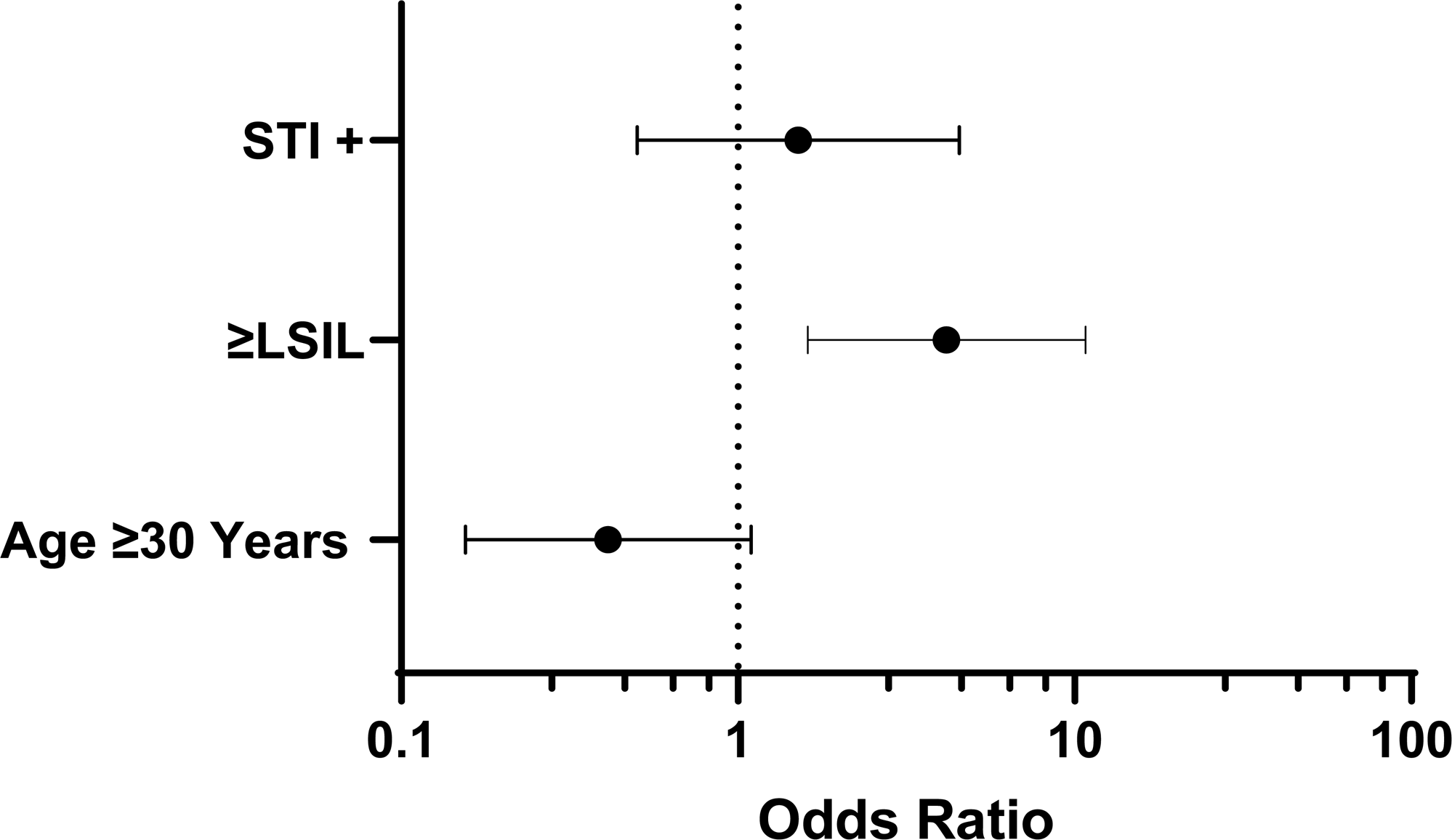
Odds ratios for factors predicting a positive Alinity m HR HPV result.

## DISCUSSION

The aim of this cohort study was to assess the prevalence and distribution of HR HPV and STI co-infection in LBC cervical specimens across cytological categories and age groups.

Recent studies have suggested a possible link between STI co-infections, HPV infection persistence, and cervical neoplasia progression (9, 10). Overall, 11.4% of the specimens were positive for a STI in this cohort and the majority (47%; n=8/17) of specimens were positive for CT. Persistent HPV and CT co-infections have been proposed as a cofactor with increased risk of progression of cervical neoplasia and malignancy in women (11). There were no NG-positive results in this study; this is consistent with the literature, where reports of an association between NG and persistent HPV infection are minimal. Globally, TV has a substantial impact on women as the most common non-viral STI and was the second most frequently observed non-HPV STI in this cohort; 66.6% (n=4/6) of specimens with a positive TV result were also positive for HPV (3). The interaction between TV infection and cervical cytological abnormalities has not been completely established; however, study outcomes have suggested that TV, though it is a predominantly asymptomatic infection, may predispose the cervical epithelium to carcinogenesis (12). MG was detected in 3 specimens: 1 was positive for HPV 16, 1 for Other HR HPV A, and 1 for Other HR HPV B. The overall prevalence of MG infection in this cohort was 2.0% (n=3/149); this is in alignment with what has been previously reported in the southern region of the United States (13). MG has emerged as a STI with a notable ability to establish chronic infections of the lower genital tract that are linked to cervical inflammation. The inflammatory response to MG infection may contribute to persistent HPV infection and an increased risk of progression to cervical disease (4, 8). The immunological effects of HPV infection or other opportunistic STI infection may have an impact on an individual’s susceptibility to other opportunistic or fastidious transmissible STI pathogens and may impair the ability to clear a persistent HPV infection from the lower genital tract.

For specimens included in this cohort, the prevalence of a progressively abnormal cytology result was higher in those with both a positive HPV and positive STI result compared with a HPV positive result only (see Figure 1). For specimens with NILM cytology, 44.6% (n=25/56) had a positive HPV result and 1.8% (n=1/56) were positive with an HPV and STI result. In specimens with ≥LSIL cytology, a near 2-fold increase in prevalence was observed in specimens with a positive HPV result (83.8%; n=31/37) and a significant increase was seen in specimens with a positive HPV and STI result (13.5%; n=5/37). The odds ratio calculations also corroborate with these observations, revealing that ≥LSIL cytology and a positive STI result were more strongly associated with a positive HR HPV result than age ≥30 years (see Figure 2). Our findings align with similar HPV/STI positivity trends associated with abnormal cytological results that have been reported in literature (14).

Our study included extended genotyping results from the Alinity m HR HPV assay (see online supplemental table 1). An Alinity m HPV16 result was associated with 1 TV (HSIL cytology) and 1 MG (ASC-US cytology) result from the Alinity m STI assay. Alinity m Other HR HPV A (31/33/52/58) result was associated with one CT (ASC-US cytology) and one MG (LSIL cytology) result. An Alinity m Other HR HPV B (35/39/51/56/59/66/68) result was linked to 4 CT, 3 TV, and 1 MG Alinity m STI results. For the Alinity Other HR HPV B results, one specimen was associated with NILM cytology, 4 with ASC-US, and 3 with ≥LSIL.

No STI co-infections with HR HPV18 or 45 were reported. All Alinity m HR HPV positive results associated with an Alinity m STI positive result were identified as a single assay response, ie, positive for HPV16, 18, 45, Other HR HPV A, or Other HR HPV B. As persistent HR HPV infections have been linked with STI co-infections, the unique extended genotyping results from the Alinity m HR HPV assay offer additional granularity to determine the causative HPV genotype risk profile for persistent infection and abnormal cytology in individuals with STI co-infections.

A positive Alinity m HR HPV and STI assay result was observed in 8.1% of specimens, with the majority having abnormal cytology (≥ASC-US). Although there appeared to be some concordance between the pathogens detected with the Alinity m STI assay and an Alinity m HR HPV result in this study, the magnitude of the potential synergistic effect between the infections on cervical disease remains unknown. The relationship between these infections can be affected by several variables including the order of infections, multiple infections across the timeline of initial infection, an inflammatory environment in the lower genital tract, and the duration of inflammation, as well as a potential genetic predisposition to cervical carcinogenesis (14).

This study has some limitations with its cross-sectional observational design. The sample size is also relatively small, symptomatic status was unknown, and histological outcomes were not available for specimens with ASC-US and ≥LSIL cytology.

In conclusion, our study revealed an association between co-infection with HPV and STI pathogens and abnormal cytology in cervical specimens. Further studies with larger sample sizes and extended HPV genotyping are needed to examine the absolute risk of cervical disease associated with co-infections by specific HPV genotypes and STI pathogens.

## Data Availability

All data produced in the present study are available upon reasonable request to the authors

## Abbreviations

AGC: atypical glandular cells
ASC-US: atypical squamous cells of undetermined significance
ASC-H: Atypical squamous cells, cannot exclude HSIL
CC: cellular control
CN: cycle number
CT: Chlamydia trachomatis
HPV: human papilloma virus
HR: high risk
HSIL: high-grade squamous intraepithelial lesion
HSV-2: Herpes Simplex Virus type 2
LBC: liquid-based cytology
LSIL: low-grade squamous intraepithelial lesion
MG: Mycoplasma genitalium
NG: Neisseria gonorrhea
NILM: negative for intraepithelial lesions or malignancy
STI: sexually transmitted infection
TV: Trichomonas vaginalis
OR: Odds Ratio

## Acknowledgements

The authors would like to thank Yan Zhang, PhD, for assistance with the statistical analysis. Stacey Tobin, PhD, provided editorial support for manuscript preparation, with compensation from Molecular Diagnostics of Abbott.

## Author contributions

Joshua Kostera, PhD: conceptualization, formal analysis, funding acquisition, methodology, supervision, writing (original draft, review & editing)

Almedina Tursunovic, MLS (ASCP): data curation, methodology

Paige Botts, MB (ASCP): data curation, methodology

April Davis, MLS (AMT): data curation

Regina Galloway, MT (ASCP): data curation

Tong Yang, MD: conceptualization, data curation, formal analysis, funding acquisition, investigation, methodology, supervision, validation, writing (review & editing)

All authors reviewed the draft and approved the manuscript for submission.

## Supplemental Materials

**Supplemental Table 1.** Distribution of Cytological Findings, by Age Group (N=149)

|  | NILM |  | ASC-US |  | ≥LSIL |  | Total |  |
| --- | --- | --- | --- | --- | --- | --- | --- | --- |
|  | n | (%) | n | (%) | n | (%) | n | (%) |
| Total | 27 | (37.6) | 69 | (37.6) | 53 | (24.8) | 149 | (100.0) |
| Age 21-29 years | 3 | (2.0) | 30 | (20.1) | 23 | (15.4) | 56 | (37.6) |
| Age 30-44 years | 21 | (14.1) | 20 | (13.4) | 15 | (10.1) | 56 | (37.6) |
| Age ≥45 years | 3 | (2.0) | 19 | (12.8) | 15 | (10.1) | 37 | (24.8) |

ASC-US, atypical squamous cells of undetermined significance; LSIL, low-grade intraepithelial lesion; NILM, negative for intraepithelial lesions or malignancy.

**Supplemental Table 2.**
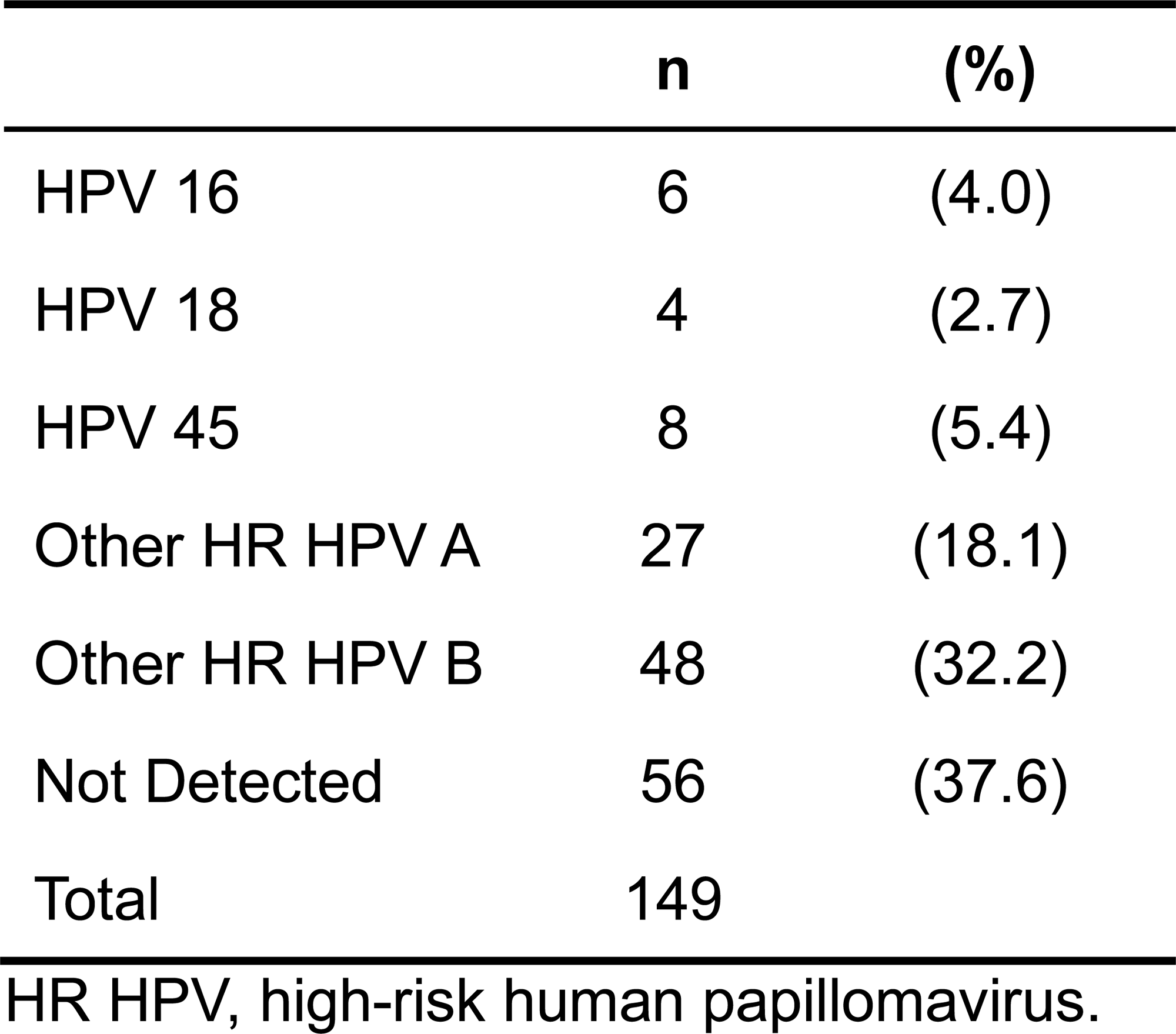
Alinity m HR HPV Result Hierarchical Distribution.

|  | n | (%) |
| --- | --- | --- |
| HPV 16 | 6 | (4.0) |
| HPV 18 | 4 | (2.7) |
| HPV 45 | 8 | (5.4) |
| Other HR HPV A | 27 | (18.1) |
| Other HR HPV B | 48 | (32.2) |
| Not Detected | 56 | (37.6) |
| Total | 149 |  |

HR HPV, high-risk human papillomavirus.

**Supplemental Table 3.** Distribution of Alinity m STI Results for each STI Pathogen, by Alinity m HR HPV Extended Genotype Result and Cytology (N=17)

|  | CT |  | NG |  | TV |  | MG |  |
| --- | --- | --- | --- | --- | --- | --- | --- | --- |
|  | n | (%) | n | (%) | n | (%) | n | (%) |
| Total | 8 | (47.0%) | 0 | (0%) | 6 | (35.0%) | 3 | (17.6%) |
| HPV16 | 0 | (0%) | 0 | (0%) | 1 | (5.9%) | 1 | (5.9%) |
| HPV18 | 0 | (0%) | 0 | (0%) | 0 | (0%) | 0 | (0%) |
| HPV45 | 0 | (0%) | 0 | (0%) | 0 | (0%) | 0 | (0%) |
| Other HR HPV A | 1 | (5.9%) | 0 | (0%) | 0 | (0%) | 1 | (5.9%) |
| Other HR HPV B | 4 | (23.5%) | 0 | (0%) | 3 | (17.6%) | 1 | (5.9%) |
| Not Detected | 3 | (17.6%) | 0 | (0%) | 2 | (11.8%) | 0 | (0%) |
| NILM | 1 | (5.9%) | 0 | (0%) | 0 | (0%) | 0 | (0%) |
| ASC-US | 6 | (35.3%) | 0 | (0%) | 3 | (17.6%) | 2 | (11.8%) |
| ≥LSIL | 1 | (5.9%) | 0 | (0%) | 3 | (17.6%) | 1 | (5.9%) |

ASC-US, atypical squamous cells of undetermined significance; CT, Chlamydia trachomatis; HR HPV, high-risk human papillomavirus; LSIL, low-grade intraepithelial lesion; MG, Mycoplasma genitalium; NG, Neisseria gonorrhea; NILM, negative for intraepithelial lesions or malignancy; TV, Trichomonas vaginalis.

